# Saliva cytokines in adult patients with different tooth caries intensity

**DOI:** 10.1101/2025.11.16.25340368

**Authors:** A.A. Bayakhmetova, A.O. Seydekhanova, A.N. Primbayeva, A.M. Duisenbayev

## Abstract

**Aim:** To study the saliva content and correlation with the intensity of caries of proinflammatory cytokines IL-1ß, IL-6, IL-8 and anti-inflammatory cytokines IL-4 and IL-10.

**Methodology:** 33 patients aged 18 to 30 years were divided into 3 groups - a group of patients without caries, a group with compensated caries at the index value the intensity of caries of the DMF (decay/ missing/ filled) is less than 5.0 and the group with decompensated caries with a DMF index value of more than 5.0.

**Results:** It was found that both compensated and decompensated caries statistically significantly reduced the concentration of anti-inflammatory cytokine IL-4. With compensated caries, a significant decrease in the content of pro-inflammatory cytokine IL-8 had a direct positive correlation with the concentration of anti-inflammatory cytokine IL-10 in saliva. The DMF index was in negative correlation with the concentration of IL-6 in saliva. With decompensated caries, there was a significant increase in the content of the proinflammatory cytokine IL-1ß in saliva, which exceeded the same indicator in other groups by 3 or more times.

**Conclusion:** The concentration of pro-inflammatory cytokines IL-6 and IL-8 in saliva had a pronounced postitive correlation with the concentration of anti-inflammatory cytokine IL-10.

## Introduction

Caries is a multifactorial infectious disease associated with the resident flora of bacterial biofilm in the oral cavity. According to its parameters, the oral cavity is characterized by extremely favorable conditions for the vital activity of the resident bacterial microflora [1,2,3,4]. In the pathogenesis of caries, an important role belongs to the immunobiological state of the patient’s body with caries and the peculiarities of the interaction of the body’s immune system and the cariesogenic microflora. Factors of local immunity of the oral cavity deserve special attention. Local protection in the oral cavity is represented by secretory immunoglobulin A (sIgA), antimicrobial peptides and a variety of other cellular and humoral factors of nonspecific and specific immunity that ensure caries resistance of hard tooth tissues and dynamic balance of de- and remineralization processes in enamel [5,6,7,8,9,10]. The object of research to assess the state of local immunity in the oral cavity is usually saliva, oral and gingival fluid, which is quite simple to obtain for research. A number of indicators – biomarkers of pathological processes for personalized monitoring of caries and other oral diseases have been identified in saliva [11,12,13].

Polypeptide molecules - cytokines, which are referred to mediators or regulators of cellular and humoral immunity in the body, take part in the formation and regulation of the protective functions of the body. Synthesis of most cytokines is induced in response to microbial invasion, antigenic irritation or tissue damage. The efficiency of cytokine synthesis in response to antigenic irritation and their activity in extremely low concentrations is similar to hormones. The high efficiency and reliability of the biological action of cytokines is ensured by their polyfunctionality or pleiotropy, which is expressed in their multifaceted activity and ability to affect cells of various types [14,15,16].

Cytokines are synthesized mainly by lymphocytes, but monocytes, macrophages, granulocytes and other cells can also be their source. According to the mechanism of action, cytokines are distinguished, providing activation (pro-inflammatory) and suppression (anti-inflammatory) of the inflammatory response, as well as regulating cellular and humoral immunity. Pro-inflammatory cytokines include interleukins 1,2,6,8, TNF-α, interferon-γ, anti-inflammatory cytokines - interleukins 4,10, TGF-ß. According to some authors, cytokines play a decisive role in the nature of the emerging adaptive immune response [17,18]. Local protective immune responses in the oral cavity, such as phagocyte chemotaxis, phagocytosis and antibody formation, are regulated by cytokines. Studies of the cytokine content in the mixed unstimulated saliva of children and adolescents with caries show a statistically significant increase in the level of proinflammatory cytokines IL-6, IL-8 and TNF-α in comparison with similar indicators of healthy people [19,20,21]. However, it should be emphasized that studies of the cytokine profile of saliva mainly concern children with caries. Studies of the imbalance of the local cytokine profile in adult patients with caries are isolated. In adult patients with caries, the concentration of sIgA in the oral fluid and IFN-γ decreased, which occurred against the background of a significant increase in the level of the proinflammatory cytokine IL-1ß [22].

The role of proinflammatory cytokines in the pathogenesis of caries in both children and adults is not sufficiently clear. It is known that they increase the permeability of the tissues of the oral cavity, which probably leads to increased caries susceptibility of the hard tissues of the tooth to the effects of cariogenic factors. Analysis of the literature data suggests age-related features of the cytokine spectrum of saliva and the nature of adaptive local immunity in adult patients with caries, which was the reason for this study.

## Materials and methods

A survey was conducted of 33 people (16 men and 17 women) aged 18 to 20 years (13) and 20-29 years (20) with medium caries. The entry criteria were the absence of somatic pathology and age up to 30 years. Three groups were formed depending on the DMF indicator. The groups with medium caries were represented by 15 patients with compensated caries with a DMF of no more than 5.0, the group with decompensated caries consisted of 18 people with a DMF of more than 5.0. The comparison group consisted of 10 practically healthy people under the age of 30 and with no caries. All patients gave informed consent to the examination and treatment of caries. In all examined patients on an empty stomach, mixed unstimulated saliva was collected by spitting into a sterile tube for 10 minutes in the morning from 9 to 11 o’clock. Saliva was centrifuged at 10000 r.p.m, the prepared samples were stored at a temperature of – 20□. Taking into account the literature data mentioned above, pro-inflammatory cytokines Il-1ß, Il-6, Il-8 and anti-inflammatory cytokines Il-4 and Il-10 were selected for the study. To determine the level of cytokines in saliva samples, a “sandwich” version of solid-phase enzyme immunoassay was used using mono- and polyclonal antibodies to cytokines Il-1ß, Il-4, Il-6, Il-8 and Il-10 and using a set of reagents and instructions from Vector Best (Russia). Sandwich-ELISA, consists in the following: one type of monoclonal antibodies to a certain cytokine is immobilized on the inner surface of the cells of the tablets for research. The test material and the corresponding standards and controls are introduced into the wells of the tablet. After incubation and washing, the second monoclonal antibodies to another epitope of this cytokine, conjugated with an indicator enzyme (horseradish peroxidase), are introduced into the wells. After incubation and washing, a substrate is introduced into the cells-hydrogen peroxide with chromogen. During the enzymatic reaction, the intensity of the color of the wells changes, which is measured on an automatic photometer for tablets. Statistical analysis of the obtained digital material was carried out in the SPSS Statistics version 22 program using descriptive statistics, the Mann-Whitney criterion for comparing the results obtained in groups and correlation analysis with the determination of Spearman’s rank correlation coefficient.

## RESULTS

The results obtained are shown in Tables 1,2 and Figures 1,2.

**Table 1.**
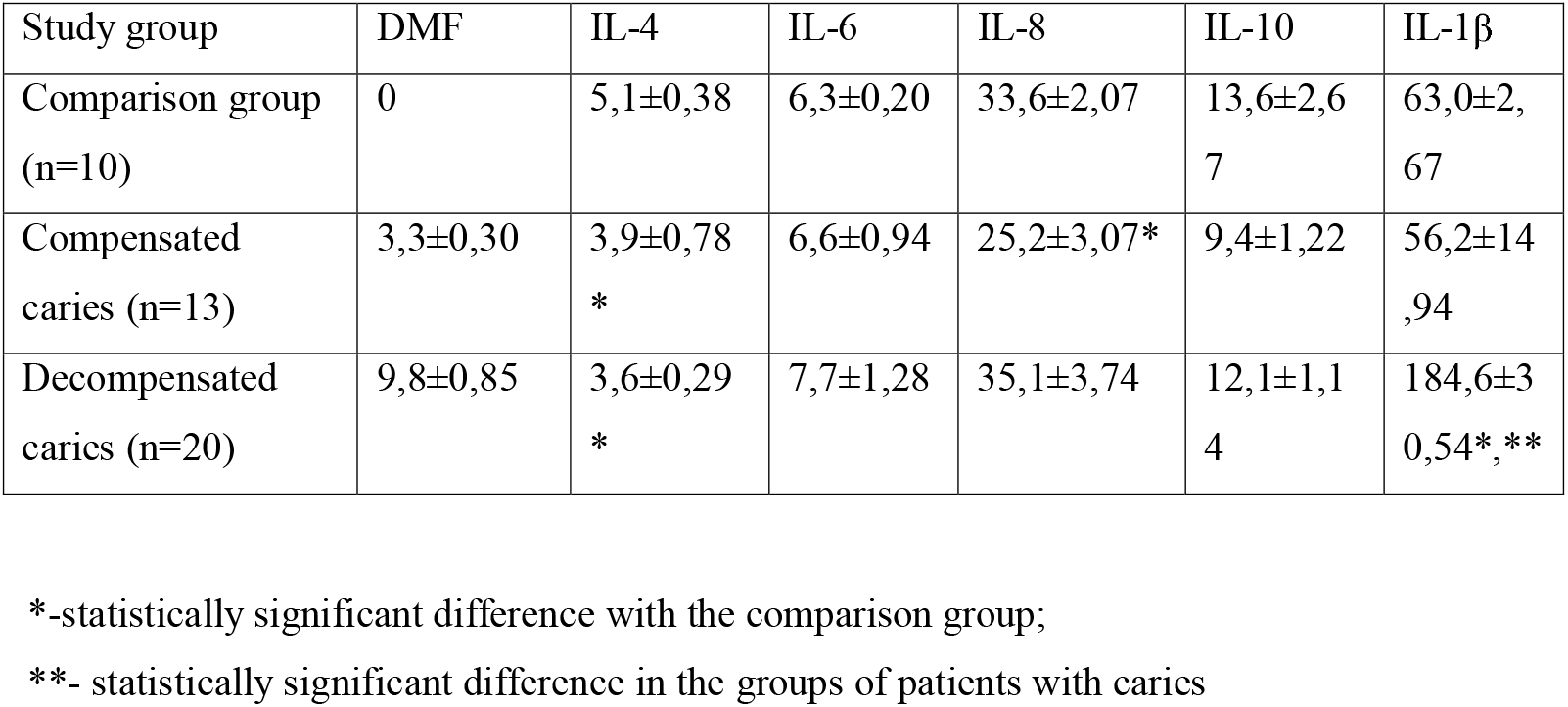
The content of cytokines in the saliva of patients with different intensity of caries (pg / l).

**Table 2.**
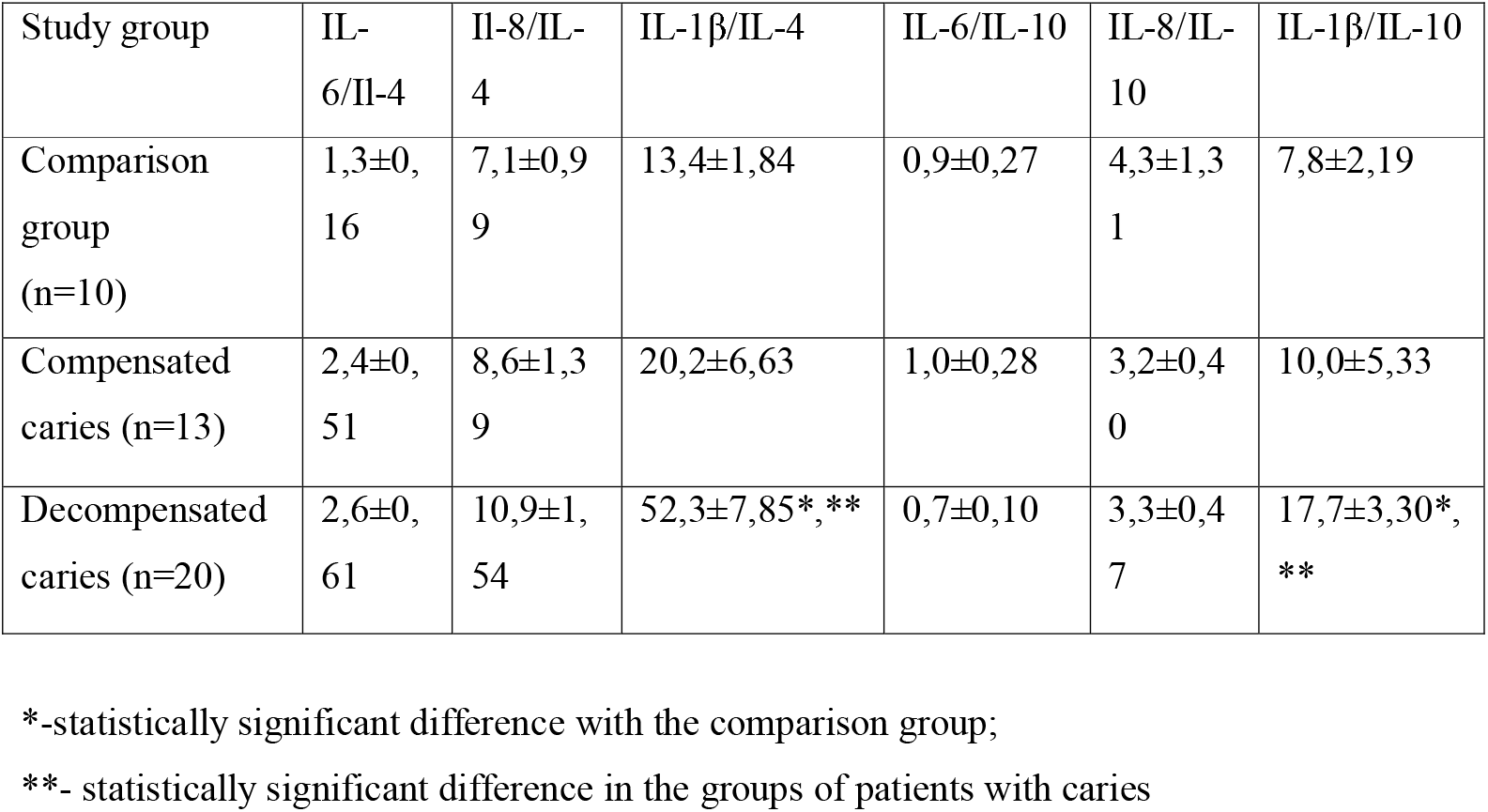
Ratios of pro- and anti-inflammatory cytokines in saliva of patients with different intensity of caries.

**Figure 1.**
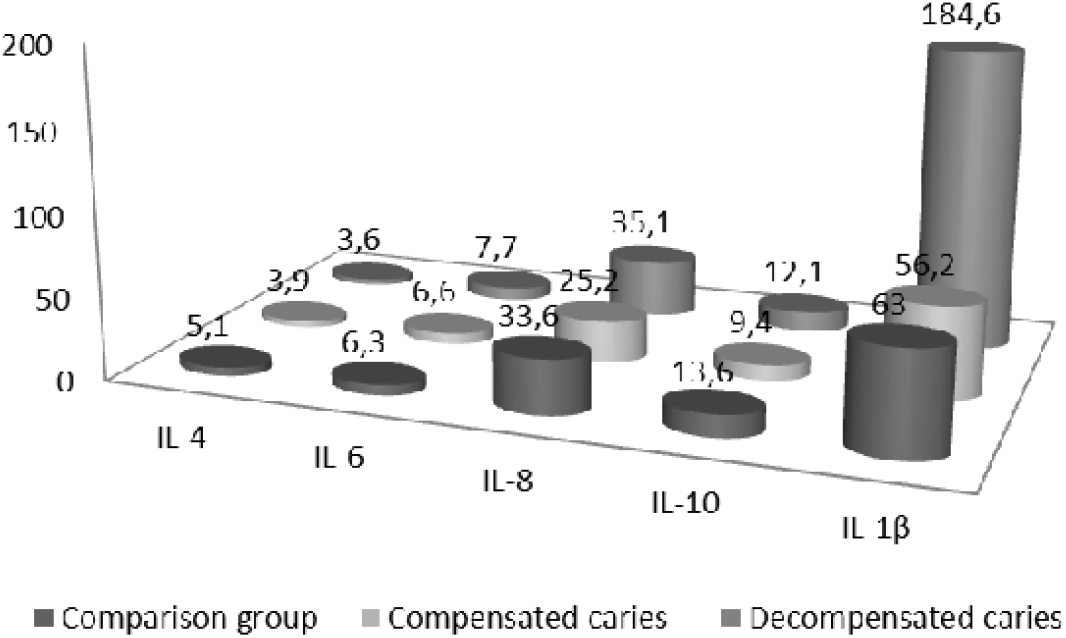
Cytokine content in saliva of patients with different intensity of caries.

**Figure 2.**
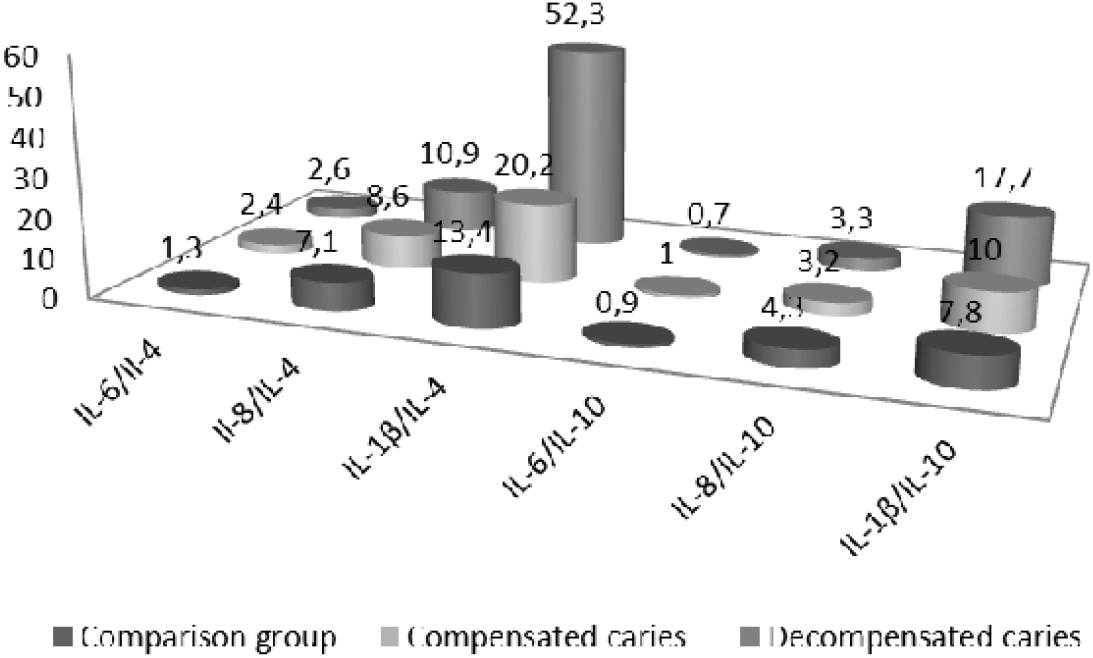
Ratios of pro- and anti-inflammatory cytokines in saliva of patients with different intensity of caries.

In the saliva of patients with compensated caries, the content of IL-4 and IL-8 decreased, this decrease was significant according to the Mann-Whitney U test (P<0.02, P<0.05, respectively). The DMF index in accordance with Spearman’s rank correlation coefficient had a significant inverse correlation with the concentration of proinflammatory cytokine IL-6 in saliva (r= -0.69, P<0.004). A positive correlation was found between the indicators of the content of IL-8 and IL-10 in saliva (r=0.64, P <0.01).

With decompensated caries, a significant decrease in the concentration of Il-4 in saliva also persisted (P<0.003), significant changes occurred with the content of Il-1ß in saliva, which increased by 2.9 and 3.3 times relative to the indicator of the comparison group and persons with compensated caries (P<0.03, P<0.001).In saliva, the concentration of pro-inflammatory cytokines IL-6 and IL-8 had a pronounced direct correlation with the concentration of anti-inflammatory cytokine Il-10 (p= 0.55, P<0.02, P=0.53, P< 0.02).

To clarify the nature of the relationship between pro- and anti-inflammatory cytokines, a study was conducted of the ratios between the content of pro-inflammatory cytokines I□-1β, I□-6, I□-8 and the concentration of anti-inflammatory cytokines I□-4 and I□-10. By the nature of the ratios, it is possible to judge the relative deficiency or hyperproduction of a particular cytokine. The results obtained are shown in table 2 and in figure 2.

As shown in Table 2, changes in the ratio of pro-inflammatory and anti-inflammatory cytokines in the saliva of patients with dentin caries reflected a significant increase in the concentration of pro-inflammatory interleukin IL-1ß, which in the group of patients with decompensated caries exceeded the indicator of the comparison group and those with compensated caries. In the group of patients with decompensated caries, the ratio of IL-1ß/IL-4 was 3.9 times higher than in the comparison group and 2.6 times higher than in the group with compensated caries (P<0.0001, P<0,001). Similar changes also affected the ratio of IL-1ß/IL-10, which was 2.3 times higher than in the comparison group and 1.8 times higher than in the group with compensated caries (P<0.05, P<0.005).

## DISCUSSION

It is known that a complex of various factors plays a role in the etiopathogenesis of caries, of which microbial biofilm, a high-carbohydrate diet of soft and sticky consistency, as well as a low level of oral hygiene were singled out as the primary ones. Caries refers to infectious diseases, in the pathogenesis of which, as is known, an important place belongs to the immune system, the state of nonspecific and specific immunity in the oral cavity. The occurrence and development of caries is reasonably associated with defects in local immunity [25,26]. The local immunity of the oral cavity, being an integral and subordinate part of the general immunity, has some features. The synthesis of protective factors occurs in the oral cavity, their content does not correlate with blood parameters, which indicates in favor of the autonomy of local immunity. The oral cavity protection system is an optimal combination of a variety of non-specific and specific protective factors that provide effective protection against cariogenic factors of microbial biofilm [27]. Saliva plays an important role in maintaining balance and symbiotic relationships between the body and the oral microbiota.

The structural features of enamel provide it with extreme strength and mechanical stability, enamel has micropores between crystals in enamel prisms, interprismatic spaces, enamel fluid is present and moves, which plays an important role both in the process of demineralization and remineralization [23,24]. The permeability of tooth enamel is determined by age, its maturity, the presence in saliva of those or other enzymes, saliva viscosity and its acidity. The group of teeth is important, as well as the localization of the enamel area.

With age, structural and functional changes occur in the hard tissues of the tooth, and the state of local immunity also becomes different. The ratio of organic and inorganic components of enamel changes, there is a decrease in organic compounds in enamel with an increase in calcium, phosphorus, zinc and fluorine. The permeability of enamel to water, ions, enzymes and amino acids decreases, there is an accumulation of minerals with a significant decrease in microspaces and micropores [25,26]. In the age groups after 35 years of age, low compliance of tooth enamel to the action of acids (<40%) with its high remineralizing ability (1-3 days) is much less common than in young people, which worsens with increasing age. A more characteristic feature of age-related enamel is its delayed remineralization (4 days or more).

Cytokine imbalance in saliva is given a determining role in the pathogenesis of caries [13]. Some pro-inflammatory and anti-inflammatory cytokines (IL-6, IL-8, TNF-α, IL-1ß, IL-4, IL-10) are associated with the pathogenesis of caries. Cytokine IL-4 is synthesized by mast cells, T-helper cells of the second type, eosinophils and basophils. IL-4 is an anti-inflammatory cytokine, stimulates the formation of reparative macrophages of the second type, promotes the differentiation of naive T-lymphocytes into T-helper cells of the second type and their transformation into plasma cells, which leads to a humoral pathway of immunity with the synthesis of specific antibodies. The anti-inflammatory cytokine IL-10 is produced by macrophages and regulatory T cells, inhibits the functions of monocytes/macrophages, the secretion of pro-inflammatory cytokines IL-1,6,8,12, TNF-α, IFN-γ by various cells. The lack of anti-inflammatory cytokine IL-10 indicates a lack of nonspecific immunity, pro-inflammatory cytokine IL-8 to inhibit phagocytic activity with impaired chemokinesis, but an adequate assessment of the cytokine profile can be carried out with mandatory consideration of the ratios of pro- and anti-inflammatory cytokines in a complex interconnected cytokine network in which the secretion of one cytokine leads to the appearance and activation of others [12]. Cytokine IL-6 is a pro- and anti-inflammatory cytokine that is synthesized by macrophages, monocytes, fibroblasts and endothelial cells. It can inhibit the synthesis of activated macrophages and T-lymphocytes of the proinflammatory cytokine TNF-α and IL-1ß. A correlation was established between the increased level of IL-6 in saliva and the intensity of caries. IL-1ß is a multifunctional cytokine with a wide spectrum of action, synthesized by macrophages and monocytes, plays a key role in the development and regulation of nonspecific protection and specific immunity, one of the first to be included in the body’s protective response under the action of pathogenic factors.The results obtained by us show that with compensated and decompensated caries in adult patients, due to a significant decrease in the content of IL-4 in saliva, violations of the formation of adaptive local immunity of the humoral type are likely. A decrease in the saliva concentration of patients with compensated caries IL-8 leads to an imbalance of nonspecific immunity, insufficiency of phagocytic activity of phagocytic cells. The inverse correlation of the IL-6 content with the DMF index in compensated caries suggests its inhibitory effect on the synthesis of the proinflammatory cytokine IL-1ß. In decompensated caries in adult patients, the system of nonspecific immunity experiences significant stress with inadequate enhancement of IL-1ß synthesis.

## CONCLUSION

The study of the cytokine profile of saliva in adult patients with medium caries and the results obtained allowed us to come to the following conclusions.

Adult patients with dentin caries have age-related features of both local immunity indicators and morphofunctional properties of hard tooth tissues. In adult patients with caries, there is a pronounced tension in the system of nonspecific immunity, which is aggravated with an increase in the intensity of caries. The imbalance of the cytokine profile of saliva in adult patients with caries indicates violations of the formation of adaptive local immunity according to the humoral type, which is noted both in patients with compensated and decompensated caries. Our results and conclusions are based on a small number of observations and require further research.

## Data Availability

All data produced in the present study are available upon reasonable request to the authors

